# Med-SSFWT: A Self-supervised Federated Weight Transfer Framework for Medical Model Fusion

**DOI:** 10.64898/2025.12.08.25340199

**Authors:** Qihan Huang, Yanju Huang, Kaijiong Zhang, Rui Yuan, Zhanyu Zhang, Yu Xiang, Zhengnan Wang, Serda Zita Milendz Ikapi, Illich Manfred Mombo, Yongzhao Zhang, Qiming Tang, Qun Yi, Haohan Zhang, Dongsheng Wang, Xiaowei Mao

## Abstract

Artificial Intelligence (AI) holds great potential to revolutionize healthcare by integrating and analyzing diverse multi-source medical data to drive advancements in disease diagnosis, treatment strategies, and patient management. However, deploying AI in distributed medical environments presents critical challenges, including data silos, label deficiency, and data heterogeneity. To address these challenges and enable effective and privacy-preserving distributed medical AI models, we propose Med-SSFWT, a Self-Supervised Federated Weight Transfer framework designed for medical data fusion. Firstly, Med-SSFWT employs a fine-tuned Large Language Model (LLM) to extract structured features from each client’s medical data, followed by feature alignment across clients via a shared global schema. Subsequently, an information gain-based gradient filtering mechanism is introduced to federated aggregation by filtering out ineffective gradients, thereby improving the robustness of global model. Furthermore, Med-SSFWT leverages a novel federated model fusion frame, consisting of self-supervised pre-training and fine-tuning through weight transfer to balance global optimization with client-specific personalization. Finally, extensive experiments show that Med-SSFWT consistently outperforms federated learning approaches in both performance and adaptability under diverse non-IID conditions, highlighting its effectiveness within distributed medical environments and establishing a foundation for the development of privacy-preserving and scalable AI-driven healthcare solutions.

## Introduction

The application of AI in healthcare has shown transformative potential, improving disease diagnosis, optimizing treatment strategies, and enhancing patient management^1–3^. AI-driven models, such as those used for predicting tumor stages in esophageal cancer at early stages, have demonstrated their ability to enable precise clinical decision-making and support the strategies of personalized treatment^4–7^. These advancements are driven by the integration of multi-source data fusion, encompassing information obtained across diverse institutions, clinical departments, and data acquisition pipelines^8–10^. This approach provides holistic insights, improves diagnostic accuracy, and enhances the reliability of healthcare systems by leveraging the strengths of multiple data modalities. Deploying AI in distributed medical environments presents unique complexities due to the fragmentation of healthcare data across institutions, heterogeneous infrastructures, and diverse data formats. Privacy regulations, such as the General Data Protection Regulation (GDPR)^11^, further constrain data sharing, making traditional centralized approaches infeasible. These factors demand innovative frameworks capable of integrating multi-source data while preserving privacy.

Navigating these complexities requires addressing several critical challenges. These challenges are pivotal to achieving effective medical data fusion and advancing the development of robust AI models. The challenges are illustrated in Fig. 1a and are as follows: (1) Healthcare data is often isolated across institutions due to stringent privacy regulations and concerns about data sharing. This isolation results in valuable medical information remaining locked within individual organizations, hindering the integration required to train robust and generalized models^12,13^. (2) Many medical datasets suffer from insufficient or inconsistent labeling, primarily due to the labor-intensive and time-consuming nature of manual annotation by domain experts^14^. In tasks such as disease stage prediction for esophageal cancer, the absence of comprehensive and consistent labels can significantly hinder model performance, as robust training critically depends on the availability of high-quality annotated datasets. (3) Medical data is inherently heterogeneous, comprising structured formats such as electronic health records (EHRs), which integrate information from diverse clinical sources, and unstructured formats including medical images and time-series signals. The non-independent and identically distributed (non-IID) nature of this data across institutions^15^, combined with variations in sensor modalities, population demographics, disease prevalence, and diagnostic protocols, further complicates achieving model generalization and effective integration.

**Figure 1.**
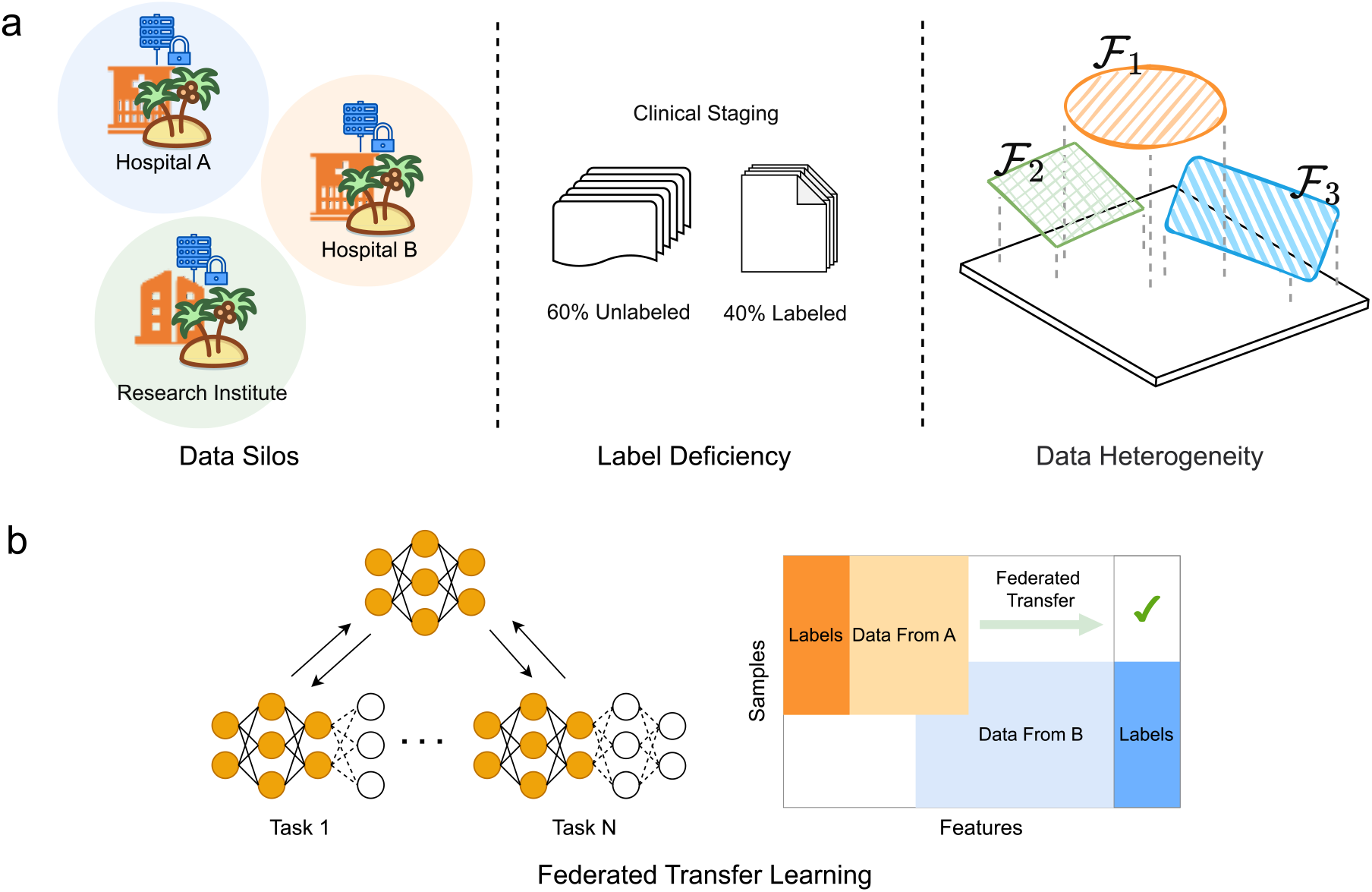
Challenges in medical data fusion and the role of federated transfer learning (FTL). (a) Major challenges: data silos, label deficiency, and data heterogeneity. (b) FTL addresses minimal overlap in feature and sample spaces across institutions, enabling collaborative training while preserving data privacy.

Addressing the challenges of deploying AI in distributed medical environments requires innovative approaches, with Federated Learning (FL) emerging as a key solution^16–19^. FL has become as a promising paradigm to enable collaborative model training across institutions without exposing raw data, thus ensuring compliance with privacy regulations while alleviating data-sharing concerns^20–22^. Techniques such as differential privacy, secure aggregation, and model personalization have further improved the scalability of FL^23^. However, traditional FL still suffers from unstable convergence and limited generalization under highly heterogeneous and non-IID medical data distributions, as well as from its strong reliance on label completeness and inefficient parameter synchronization. To mitigate these issues, Federated Transfer Learning (FTL) extends FL by incorporating pre-trained models and transfer mechanisms to adapt to domain shifts and label deficiencies^22,24^ (Fig. 1b). Yet, most FTL frameworks still perform parameter-level aggregation, which fails to handle semantic misalignment across clients and often causes performance degradation in cross-institutional settings^25–27^. In parallel, Large Language Models (LLMs) have recently shown great promise in medical text understanding and structured feature extraction, offering a pathway to reduce annotation costs and enhance model generalizability^28^. Nevertheless, their integration into federated or distributed frameworks remains challenging due to communication overhead, privacy risks during fine-tuning, and the absence of a unified representation alignment strategy.

To overcome the limitations of existing FTL frameworks in distributed medical AI, we propose Med-SSFWT, a novel Self-Supervised Federated Weight Transfer framework. The primary contributions of this paper can be summarized as follows: (1) A personalized approach for distributed feature extraction leverages each client’s unique medical text corpus to fine-tune a LLM, enabling the generation of standardized, text-level structural features while preserving client-specific information; (2) An Information gain-based Gradient Filtering Mechanism (IGFM) is developed to optimize federated aggregation by filtering ineffective gradients based on their data contribution, enhancing the global model’s robustness and mitigating the challenges posed by data heterogeneity; (3) A novel federated model fusion approach is proposed, consisting of a pre-training FL stage that performs self-supervised learning of a representation module, followed by a fine-tuning FL stage that adapts the model to client-specific tasks achieving a balance between global optimization and personalization.

## Results

### Datasets and overall framework

The experiments employed three comprehensive datasets. The primary dataset is the Sichuan Cancer Hospital Esophageal Cancer dataset (SCCH-ESCA), which contains 6,025 records with both structured and unstructured clinical data and is used to evaluate the performance of the proposed FL model. In addition, the Cancer Genome Atlas Esophageal Cancer dataset (TCGA-ESCA) includes 3,389 records with genomic and clinical annotations, while the Surveillance, Epidemiology, and End Results Esophageal Cancer dataset (SEER-ESCA) comprises 11,921 records with detailed clinical and demographic information; both datasets are utilized to determine and pre-train the backbone model architecture. These datasets provide detailed staging information, covering tumor stages (T0–T4), lymph node involvement (N0–N4), and metastasis status (M0, M1). All esophageal cancer types were analyzed collectively to enhance statistical reliability, as squamous cell carcinoma accounted for over 96% of cases, while adenocarcinoma and adenosquamous carcinoma together comprised less than 4%.

Med-SSFWT consists of a central server coordinating multiple distributed clients without sharing raw data. Each client locally extracts structured clinical features from unstructured medical records using fine-tuned LLM. The training process consists of two stages: (1) self-supervised federated pre-training, where clients independently pre-train a shared model on their local data, and the server aggregates these updates into a global representation model; (2) federated fine-tuning, where the global model is further optimized alongside client-specific prediction heads.

### Med-SSFWT outperforms other FL algorithms

The default training parameters for our proposed model are as follows: the number of local clients *N*_client_ is set to 5, and the dataset is randomly and evenly divided among them. In each communication round, the client participation rate *Q* is set to 0.6, and the total number of global rounds is 20. The learning rate for the shared global parameters *η* is set to 2**×**10^-5^, the batch size *N*_batch_ is 64, and the Dirichlet distribution parameter α is set to 0.1. In addition, the masking ratio, used in the masked language modeling pre-training task, is set to 0.15, and the number of local epochs on each client per communication round is set to 10. These default values provide a balanced and consistent baseline for evaluating our model. To further assess the adaptability and robustness of the proposed framework under varying FL scenarios, we perform additional experiments by systematically varying key hyperparameters. Specifically, the Dirichlet distribution parameter *α* is adjusted to simulate different degrees of data heterogeneity across clients; the total number of local training epochs is varied to explore the trade-off between local computation and global communication; and the masked ratio used in self-supervised pre-training is tuned to evaluate its impact on representation quality and downstream performance.

To assess the effectiveness of the proposed framework, Med-SSFWT, several baseline FL algorithms are selected for comparison. FedAvg^29^, the most widely used baseline, updates the global model by averaging client updates. FedProx^30^ extends FedAvg by introducing a proximal term to the local objective function, which helps mitigate the impact of data heterogeneity and ensures stable convergence. FedAdam^31^ adapts the Adam optimizer to the federated setting, enabling adaptive learning rates for faster and more efficient training. FedNova^32^ tackles the challenges of local update heterogeneity caused by varying client participation and training epochs by normalizing updates before aggregation. All baseline methods, including FedProx, FedAdam, and FedNova, employ BERT as the backbone and rely on structured features, without incorporating gradient filtering. FedProx and FedAdam are both optimized using Adam, whereas FedNova adopts SGD. In contrast, the proposed Med-SSFWT directly leverages raw textual features and integrates a gradient filtering mechanism to enhance training stability and robustness, while also employing the Adam optimizer. This configuration highlights the methodological innovations of Med-SSFWT in terms of input representation and optimization strategy. In experiments, Bidirectional Encoder Representations from Transformers (BERT) was employed as the backbone architecture for FL framework. To evaluate the training efficiency and convergence of models, we introduce three convergence-related metrics. RoA@σ measures the number of global rounds required to reach a target accuracy threshold σ, reflecting convergence speed. AoR@ γ captures the highest accuracy achieved within the first γ global rounds, indicating early-stage performance. PCE evaluates model efficiency by combining convergence speed, early performance, and final accuracy.

The performance comparison of different models under varying levels of non-IID conditions are shown below: Light Non-IID (*α*=1.0), Moderate Non-IID (*α*=0.1), and Severe Non-IID (*α*=0.01). Across different non-IID conditions, Med-SSFWT consistently outperforms baseline methods in terms of RoA, AoR, and PCE. Under the Light non-IID condition, Med-SSFWT converges with the lowest RoA@60 of 1.67 ± 1.15 and achieves the highest AoR@10 of 92.20 ± 1.23, together with the best PCE@{80,10} of 3.93 ± 3.31, surpassing FedProx (AoR@10 = 85.77 ± 1.13) and FedNova (AoR@10 = 84.97 ± 1.44), while FedAdam remains ineffective with poor AoR (60.47 ± 1.94) and negligible PCE (0.18). Under the Moderate non-IID condition, Med-SSFWT maintains robustness, achieving AoR@10 of 89.14 ± 1.28 and PCE@{80,10} of 3.54 ± 3.10, whereas FedProx and FedNova exhibit substantial drops (e.g., FedProx: AoR@10 = 80.06 ± 3.36, PCE@{80,10} = 1.02 ± 0.17; FedNova: AoR@10 = 75.17 ± 1.67, PCE@{80,10} not measurable). Even in the Severe non-IID condition, Med-SSFWT sustains superior performance with AoR@10 of 86.97 ± 1.56 and PCE@{80,10} of 2.19 ± 1.42, in contrast to the sharp degradation observed for FedProx (AoR@10 = 78.70 ± 2.93, PCE@{80,10} = 0.71) and FedNova (AoR@10 = 75.49 ± 3.01, PCE@{80,10} not measurable), and the persistent failure of FedAdam (AoR@10 = 57.11 ± 0.36). These results highlight the efficiency and robustness of Med-SSFWT in achieving favorable convergence (RoA), maintaining higher accuracy levels (AoR), and ensuring stable convergence-efficiency trade-offs (PCE) across diverse non-IID environments.

A comprehensive comparison of model performances in terms of training and testing accuracy as well as F1-Score under different non-IID conditions is illustrated in Figs. 5a-b. Under varying levels of non-IID conditions, Med-SSFWT consistently outperforms baseline methods in terms of accuracy and F1-Score. In the noise-free setting, Med-SSFWT achieves the highest train and test accuracies across all distributions, showing only limited degradation as the degree of heterogeneity increases. Specifically, it attains test accuracies of 91.50 ± 0.43%, 88.12 ± 1.34%, and 86.40 ± 1.78% under Light, Moderate, and Severe Non-IID scenarios, respectively, substantially outperforming FedProx and FedNova, whereas FedAdam consistently underperforms, with accuracies below 65% and inferior F1-scores. When additional gradient noise is introduced, all baseline methods exhibit further degradation. FedAdam nearly collapses, with test accuracy consistently below 60%; for example, it achieves only 58.01±0.27% under Light non-IID and 55.99 ± 0.39% under Severe non-IID, while F1-scores remain restricted to around 0.24-0.26. FedProx and FedNova retain moderate stability, yet both experience substantial deterioration under severe non-IID, where test accuracy decreases to 71.01 ± 0.50% for FedProx and 69.77 ± 1.65% for FedNova, accompanied by F1-scores below 0.6. In contrast, Med-SSFWT demonstrates notable robustness, sustaining 83.12 ± 1.47% test accuracy and an F1-score of 0.76 ± 0.02 even in the most adverse setting. These results underscore the superior ability of Med-SSFWT to preserve convergence stability and generalization capacity under both heterogeneous data distributions and noisy gradient perturbations, substantially outperforming existing federated optimization baselines.

Med-SSFWT demonstrates consistent superiority over baseline models across varying non-IID conditions, achieving higher AoR, PCE, accuracy, and F1-Score with minimal degradation. These results highlight its robust adaptability and strong generalization ability, indicating promising applicability in heterogeneous FL scenarios.

### Evaluation of LLM-based personalized feature extraction

A comprehensive evaluation of the effect of incorporating LLM-based personalized feature extraction into the Med-SSFWT framework under diverse training configurations is presented in Fig. 5c. Across batch sizes of 16, 32, and 64 and learning rates set to 2e-4, 2e-5, and 2e-6, the Med-SSFWT w/ LLMs consistently outperforms the baseline Med-SSFWT (without LLMs). This performance advantage is evident across all metrics, including training and testing accuracy, precision, recall, and F1-score. Notably, the test F1-score increases substantially from approximately 0.22-0.33 in Med-SSFWT to 0.74-0.79 in Med-SSFWT w/ LLMs, underscoring the significant contribution of semantically enriched features to the model’s generalization ability.

Beyond predictive improvements, the LLM-augmented variant also demonstrates greater robustness and stability. Whereas the model without LLMs exhibits pronounced sensitivity to hyperparameter changes and suffers severe degradation at extreme learning rates or larger batch sizes, the full Med-SSFWT w/ LLMs maintains consistent outcomes, with test accuracies around 0.74-0.83 and test precisions/recalls exceeding 0.76-0.89 across all settings. These results suggest that LLM-based feature extraction not only enhances the expressiveness of personalized representations but also mitigates the adverse effects of suboptimal training conditions. Overall, integrating LLMs into Med-SSFWT proves to be an effective strategy for improving adaptability and resilience in FL under heterogeneous environments.

### Effectiveness of the IGFM algorithm

The robustness of the proposed IGFM algorithm within the Med-SSFWT framework was examined by perturbing gradient updates with Gaussian noise of varying intensities (σ = 1, 0.1, 0.01) to simulate noisy training environments. This design aimed to assess whether IGFM can suppress noisy gradients while retaining informative learning signals. All experiments were conducted with a masking ratio of 0.15, using 10 global rounds, 10 local epochs, a batch size of 16, and 5 clients.

Demonstrated in Fig. 6a is that Med-SSFWT with IGFM consistently outperforms its counterpart without IGFM across all noise levels. Under the high-noise setting (σ = 1), IGFM yields noticeable gains, raising the Test Accuracy from approximately 82% to 85% and the F1-Score from approximately 0.73 to 0.78. When the noise intensity decreases, the advantage becomes even more evident: at σ = 0.01, Med-SSFWT with IGFM achieves approximately 90% Test Accuracy and 0.87 F1-Score, compared to approximately 86% and 0.81 without IGFM. These results demonstrate that IGFM effectively mitigates the adverse effects of noisy gradients, thereby enhancing both convergence stability and generalization under FL with gradient perturbations.

### Optimal selection of backbone model for Med-SSFWT

Backbone architectures serve as fundamental components influencing the overall performance of FL frameworks. In this study, four backbone models, BERT^33,34^, RoBERTa^35^, XLNet^36^, and TextCNN^37^, are evaluated on the TCGA-ESCA and SEER-ESCA datasets, focusing on key metrics including accuracy, precision, recall, and F1-Score. Each model offers a unique approach to feature extraction, ranging from simple convolutional architectures to sophisticated pre-trained transformer-based frameworks. The analysis highlights notable differences in their effectiveness, showcasing the strengths and limitations of each approach.

The performance comparison, revealed in Fig. 6b, underscores the superiority of pre-trained transformer models, particularly BERT, over simpler architectures like TextCNN. While TextCNN is computationally efficient, it converges slowly and delivers significantly lower performance, with an accuracy of approximately 80% and an F1-Score below 0.6 by the end of training. In contrast, BERT achieves rapid convergence, reaching over 95% accuracy and near-perfect precision, recall, and F1-Scores within the first five epochs. This highlights the exceptional capacity of BERT for modeling bidirectional context and capturing rich semantic information. RoBERTa, as an enhanced version of BERT, also performs exceptionally well, achieving comparable metrics with slightly higher stability in some cases. XLNet, though competitive, exhibits greater variability in precision and recall due to its permutation-based training strategy.

### Assessing the effective masking ratio in pre-training

Masked Language Modeling (MLM) is a self-supervised pre-training task utilized in transformer-based models like BERT, enabling models to learn contextual representations from unlabeled data. By masking tokens in input text and predicting them, MLM helps build effective embeddings for downstream tasks. To identify an optimal masking ratio, we experimented with varying ratios during pre-training, aiming to balance model performance and generalization in subsequent fine-tuning tasks.

The effect of different masking ratios (MR) on global perplexity reduction during the self-supervised pre-training stage of the Med-SSFWT framework is illustrated in Supplementary Fig. 1. Across all settings, global perplexity decreases rapidly within the first few global rounds and stabilizes at low levels by approximately the 6th–8th round, indicating effective convergence. Lower masking ratios (MR = 0.05 and 0.10) converge more quickly and achieve lower final perplexity values, whereas higher ratios (MR = 0.20 and 0.25) converge more slowly and remain at relatively higher perplexity levels. Importantly, MR = 0.15 provides a balanced trade-off, combining efficient convergence speed with sufficiently low perplexity, which aligns with its superior downstream task performance.

The performance metrics, including training accuracy, training F1-score, test accuracy, and test F1-score, across different masking ratios are shown in Fig. 7a. Among the evaluated settings, MR = 0.15 consistently delivers the best and most stable test performance, demonstrating stronger generalization than other ratios. MR = 0.15 achieves both higher accuracy and F1-scores with reduced variance, suggesting that this ratio provides an optimal balance between effective representation learning and generalization capacity.

Thus, synthesizing insights from both figures, we conclude that a masking ratio of 0.15 achieves an optimal balance, as it maintains low perplexity during pre-training and delivers robust, generalized performance during subsequent fine-tuning and evaluation phases. This finding aligns with the widely adopted masking ratio of 0.15 utilized in traditional BERT-based models, further supporting its efficacy in capturing meaningful contextual information for downstream tasks.

### Analysis of local training epochs in fine-tuning

The impact of varying the number of local epochs (NOLE) on model performance in FL is illustrated in Fig. 7b. The results highlight that increasing NOLE generally improves performance metrics, including Accuracy, Precision, Recall, and F1-Score, but the benefits diminish as NOLE becomes larger. This indicates a trade-off between computational efficiency at the local level and communication efficiency across the global network, particularly as NOLE increases beyond a certain point.

Faster convergence and improved accuracy are led to by higher NOLE values, as illustrated in Fig. 7b(i). By the 10th global round, models with NOLE = 20 achieve accuracy levels exceeding 90%, whereas those with NOLE = 5 reach only around 70% accuracy even by the 20th round. However, the improvement between NOLE = 15 and NOLE = 20 is minimal, suggesting diminishing returns with larger NOLE values. Notably, models with NOLE = 10 exhibit a strong balance, achieving close to 90% accuracy by the 15th round and steadily converging without the need for excessive local training. This indicates that setting NOLE = 10 achieves a balance between computational efficiency and effectiveness, making it an optimal choice for practical applications.

Similar trends are observed in Precision and Recall, where NOLE = 20 yields the highest Precision (approaching 90%) and Recall (nearing 85%). However, models with NOLE = 10 achieve comparable levels, with Precision exceeding 85% and Recall nearing 80%, providing a practical trade-off between performance and training cost. Fig. 7b(iv) further illustrates that F1-Score follows a similar pattern. While NOLE = 20 achieves the highest F1-Score, nearing 90% by the 10th global round, NOLE = 10 reaches almost the same level by the 15th round, while avoiding the diminishing returns and increased computational overhead associated with higher NOLE values.

## Discussion

This study introduces Med-SSFWT, a novel self-supervised federated weight transfer framework designed to address critical challenges inherent in deploying AI within distributed medical environments, such as data silos, label deficiency, and data heterogeneity. We evaluate Med-SSFWT on comprehensive real-world datasets, with SCCH-ESCA serving as the primary dataset for model evaluation, while TCGA-ESCA and SEER-ESCA are utilized to guide the selection and pre-training of the backbone architecture. Our results demonstrate that Med-SSFWT consistently outperforms established FL algorithms (e.g., FedProx, FedAdam, and FedNova) across varying non-IID conditions. The superior performance is attributed to three primary processes: (1) personalized distributed feature extraction via client-specific LLM fine-tuning, enabling standardized structural features with local data preservation; (2) an IGFM mechanism enhancing aggregation robustness by selectively filtering ineffective gradients; and (3) federated model fusion combining self-supervised pre-training for generalized representation learning with client-specific fine-tuning, balancing global optimization and local personalization.

The superior performance of Med-SSFWT arises from the integration of three complementary processes. Client-specific LLM fine-tuning enables semantic alignment across heterogeneous medical records, IGFM selectively aggregates high-contribution updates to ensure training stability, and the federated weight transfer strategy balances global representation learning with local personalization. As evidenced in Fig. 6a, IGFM substantially improves accuracy and F1-scores under noisy gradient perturbations, underscoring its potential to safeguard FL reliability in real-world medical environments where data quality is often inconsistent.

In contrast, Med-SSFWT consistently achieves superior classification performance and faster convergence on SCCH-ESCA, as evidenced by its higher accuracy and F1-Scores across global rounds in Fig. 5a-b and Table 2. This advantage is attributed to its personalized distributed feature extraction using client-specific LLMs, which enables semantic alignment of medical records despite structural inconsistencies. Besides, IGFM ensures that only high-contribution updates are integrated into the global model, thereby enhancing training stability and mitigating the negative effects of noisy or uninformative gradients. Together, these components facilitate the learning of a globally coherent yet locally adaptable representation, leading to efficient convergence and robust performance in clinically heterogeneous federated environments.

**Table 1.**
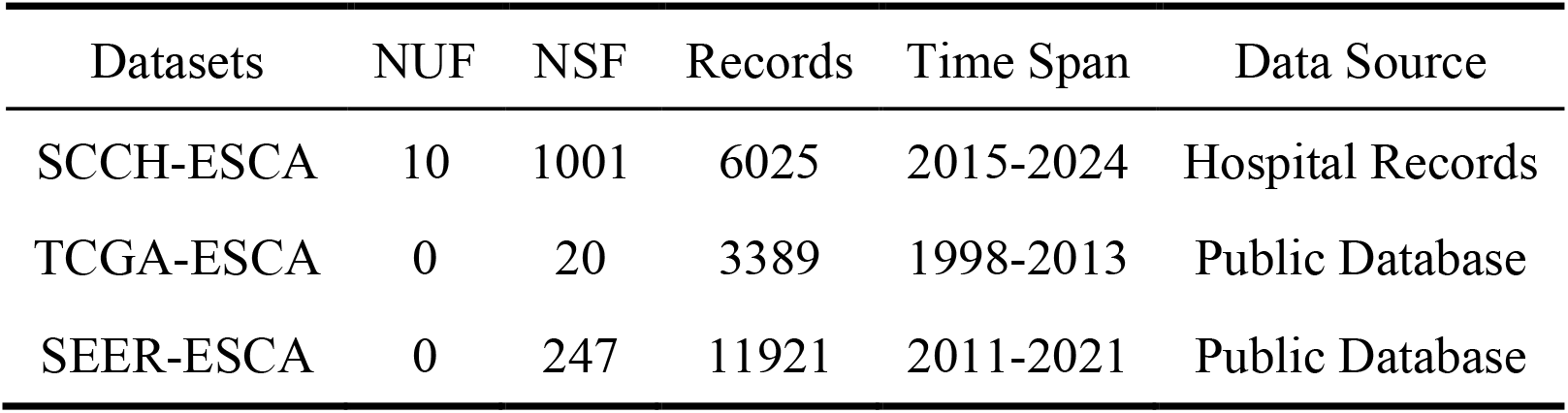
Experimental datasets include SCCH-ESCA, a hospital-based clinical record dataset from Sichuan Cancer Hospital; TCGA-ESCA, a public genomic and clinical dataset; and SEER-ESCA, a population-level registry dataset from the SEER program. Dataset characteristics are shown, including the number of unstructured features (NUF), structured features (NSF), records, time span, and data source.

**Table 2.** Comparison of convergence performance under non-IID conditions for the TNM staging of esophageal cancer. Metrics include RoA at accuracy thresholds (60%, 70%, 80%), AoR over specific epochs (5, 8, 10), and PCE combining accuracy thresholds with convergence epochs. Results are presented across three levels of non-IID data heterogeneity including Light Non-IID, Moderate Non-IID, and Severe Non-IID.

| Non-IID Degree | Models | RoA@60 | RoA@70 | RoA@80 | AoR@5 | AoR@8 | AoR@10 | PCE@<br>{60,5} | PCE@<br>{70,8} | PCE@<br>{80,10} |
| --- | --- | --- | --- | --- | --- | --- | --- | --- | --- | --- |
| Light Non-IID | FedProx | $2.00 \pm 1.73$ | $3.00 \pm 1.73$ | $3.67 \pm 2.89$ | $80.97 \pm 6.67$ | $85.18 \pm 0.98$ | $85.77 \pm 1.13$ | $4.68 \pm 2.92$ | $2.53 \pm 1.12$ | $2.43 \pm 1.34$ |
| | FedAdam | 10.00 | - | - | $56.66 \pm 1.39$ | $56.91 \pm 1.53$ | $60.47 \pm 1.94$ | 0.18 | - | - |
| | FedNova | $2.33 \pm 1.53$ | $2.67 \pm 2.08$ | $4.33 \pm 2.31$ | $79.52 \pm 5.41$ | $84.74 \pm 1.05$ | $84.97 \pm 1.44$ | $3.53 \pm 2.42$ | $3.57 \pm 2.55$ | $1.70 \pm 0.70$ |
| | Med-SSFWT | $1.67 \pm 1.15$ | $2.00 \pm 1.73$ | $3.00 \pm 2.00$ | $89.05 \pm 2.42$ | $91.54 \pm 2.17$ | $92.20 \pm 1.23$ | $5.82 \pm 2.95$ | $5.70 \pm 3.25$ | $3.93 \pm 3.31$ |
| Moderate Non-IID | FedProx | 3.00 | $4.00 \pm 1.00$ | $5.50 \pm 0.71$ | $77.06 \pm 3.50$ | $79.80 \pm 3.71$ | $80.06 \pm 3.36$ | $1.69 \pm 0.17$ | $1.38 \pm 0.45$ | $1.02 \pm 0.17$ |
| | FedAdam | 10.00 | - | - | $56.63 \pm 1.39$ | $56.90 \pm 1.53$ | $59.24 \pm 1.26$ | 0.18 | - | - |
| | FedNova | $6.67 \pm 1.53$ | $8.67 \pm 1.53$ | - | $57.49 \pm 4.74$ | $65.91 \pm 6.64$ | $75.17 \pm 1.67$ | $0.52 \pm 0.13$ | $0.45 \pm 0.10$ | - |
| | Med-SSFWT | $2.00 \pm 1.00$ | $2.00 \pm 1.00$ | $3.00 \pm 1.73$ | $87.65 \pm 1.78$ | $88.83 \pm 0.92$ | $89.14 \pm 1.28$ | $4.26 \pm 2.53$ | $4.31 \pm 2.50$ | $3.54 \pm 3.10$ |
| Severe Non-IID | FedProx | $5.00 \pm 2.65$ | $6.33 \pm 3.21$ | 8.00 | $68.80 \pm 11.72$ | $72.21 \pm 10.58$ | $78.70 \pm 2.93$ | $1.09 \pm 0.70$ | $0.88 \pm 0.51$ | 0.71 |
| | FedAdam | - | - | - | $56.63 \pm 0.13$ | $57.08 \pm 0.41$ | $57.11 \pm 0.36$ | - | - | - |
| | FedNova | $4.67 \pm 3.79$ | $8.00 \pm 1.73$ | - | $63.64 \pm 5.95$ | $67.75 \pm 8.71$ | $75.49 \pm 3.01$ | $1.21 \pm 0.82$ | $0.52 \pm 0.15$ | - |
| | Med-SSFWT | $1.67 \pm 1.15$ | $3.00 \pm 1.73$ | $4.00 \pm 2.00$ | $84.04 \pm 5.07$ | $86.31 \pm 2.20$ | $86.97 \pm 1.56$ | $5.32 \pm 2.85$ | $3.63 \pm 3.45$ | $2.19 \pm 1.42$ |

The proposed IGFM significantly enhances the resilience and generalization capability of FL by selectively aggregating informative gradient updates while filtering out noisy or low-quality signals. As shown in Fig. 6a, IGFM consistently improves both accuracy and F1-Score across varying noise levels, demonstrating its effectiveness in preserving meaningful learning signals under noisy training conditions. This selective aggregation strategy mitigates the detrimental impact of unreliable gradients and contributes to more stable convergence and improved model robustness. These results highlight the practical value of IGFM in enhancing the reliability of FL systems deployed in privacy-sensitive and data-quality-variable domains, such as healthcare.

Despite its advantages, Med-SSFWT has several limitations. First, the current evaluation focuses solely on textual data, which may not directly generalize to multi-modal clinical inputs such as imaging and laboratory results. Second, the adaptive gradient filtering mechanism, while beneficial, introduces an estimated 8–12% increase in client-side computation. Finally, the framework assumes synchronous and reliable client participation, which may not hold in real-world FL deployments. Future research will extend Med-SSFWT to multi-modal FL, explore asynchronous communication strategies, and integrate stronger privacy-preserving mechanisms such as differential privacy and secure aggregation.

## Methods

### Datasets

The Esophageal Cancer dataset from Sichuan Cancer Hospital (SCCH-ESCA) comprises 6,025 records, including 2,897 labeled samples, with both structured and unstructured clinical data. The structured data includes measurements such as blood pressure, height, weight, body surface area, and laboratory test indicators such as γ-glutamyl transferase (γ-GT) and alanine aminotransferase (ALT). The unstructured data encompasses detailed medical histories, diagnostic findings, and treatment records. The dataset follows the 8th edition TNM classification system, providing staging information for (a) Tumor stages (T0 to T4), (b) Lymph node involvement (N0 to N4), and (c) Metastasis status (M0 and M1), as illustrated in Fig. 4a.

The TCGA-ESCA dataset, derived from The Cancer Genome Atlas (TCGA), includes 3,389 records of esophageal cancer patients spanning the years 1998 to 2013. This dataset is publicly available and provides 20 structured clinical features, such as tumor stage, grade, and patient demographics. The Surveillance, Epidemiology, and End Results (SEER) program, maintained by the National Cancer Institute (NCI), provides comprehensive cancer-related data across various regions in the United States, offering valuable insights into the incidence, treatment, and survival outcomes of different cancers. The SEER-ESCA dataset is specifically extracted from the SEER program, focusing on esophageal cancer. It includes clinical and demographic data of patients diagnosed between 2011 and 2021, comprising variables such as tumor stage, histology, treatment modalities, survival rates, and demographic details.

The data analysis for esophageal cancer was conducted by including all cases as a unified cohort, without stratifying between squamous cell carcinoma and adenocarcinoma. This strategy was adopted to enlarge the effective sample size and thereby enhance statistical reliability and model robustness. Importantly, squamous cell carcinoma constituted more than 96% of the cases in the dataset, while adenocarcinoma and adenosquamous carcinoma together accounted for less than 4%. Given this distribution, unified analysis not only ensured reliable statistical power but also provided a representative assessment of the model’s applicability to esophageal cancer by integrating both structured and unstructured data sources.

### System configuration

All experiments were conducted on a high-performance computing platform. Model training and large-scale simulations were executed on a dedicated server configured with 2 NVIDIA Tesla A800 GPUs (80 GB memory per card), dual Intel Xeon Gold 6348 CPUs providing a total of 56 cores, and 503 GB of RAM, operating under a Linux-based environment. The software stack consisted of Python 3.10.14 and PyTorch 2.4.1. In addition, data preprocessing and supplementary analyses were performed on a separate workstation equipped with a 24-core CPU, an NVIDIA RTX A6000 GPU, and 128 GB of RAM.

### Problem formulation

In the training of Med-SSFWT, a two-stage process is utilized: (1) the pre-training of a global model based on BERT, and (2) the fine-tuning of the global model, which incorporates shared neural network layers along with client-specific task heads designed for various downstream applications.

The first stage involves the pre-training of a global BERT model using a self-supervised learning objective. In this stage, the global model is trained collaboratively across all clients, with the aim of learning generalizable representations from unlabeled data. The optimization objective can be defined as Eq. (1):

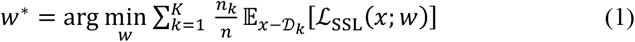

where *w* represents the global model parameters, 𝒟_*k*_ is the local data distribution of client *k*, ℒ_SSL_(*x*; *w*) is the self-supervised loss function (e.g., masked language modeling or masked token prediction), *n*_*k*_ is the number of samples at client *k*, and 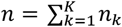 is the total number of samples across all clients.

After obtaining the pre-trained global model weights *w*^∗^, the second stage focuses on fine-tuning the model for specific downstream tasks. Each client trains a shared neural network layer based on the pre-trained global model and configures client-specific task heads tailored to its local dataset. The objective for the fine-tuning process is expressed as Eq. (2):

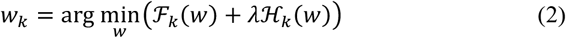

where ℱ_*k*_(*w*) represents the supervised loss on client *k* ‘s data, ℋ_*k*_(*w*) is the loss function of client-specific task head, and λ is a regularization parameter that balances the contributions of the shared neural network and client-specific components.

The shared neural network layers retain the global representations learned during the pre-training, ensuring the generalization across clients. Meanwhile, the client-specific task heads allow the model to adapt to the unique characteristics of each client’s data. This two-stage framework improves both the generalization and personalization capabilities of the FL system, effectively balancing global and local objectives.

### Architecture of Med-SSFWT

Med-SSFWT adopts an FL paradigm composed of a central server and multiple distributed clients. Instead of pooling raw data at the server, each client utilizes its local data to train a shared representation model, which includes self-supervised pre-training on text and structured clinical features, while maintaining client-specific prediction heads locally and confidentially.

The architecture of Med-SSFWT is illustrated in Fig. 2, a FL framework that integrates local feature extraction and global model fusion. The framework begins with data collection, where each client processes unstructured and structured medical records. In the personalized feature extraction module, unstructured data is transformed into structured feature sets using fine-tuned LLMs, ensuring compatibility with downstream tasks. Subsequently, each client trains a local model that combines BERT-based encoders and client-specific task heads, adapting to its unique dataset and requirements.

**Figure 2.**
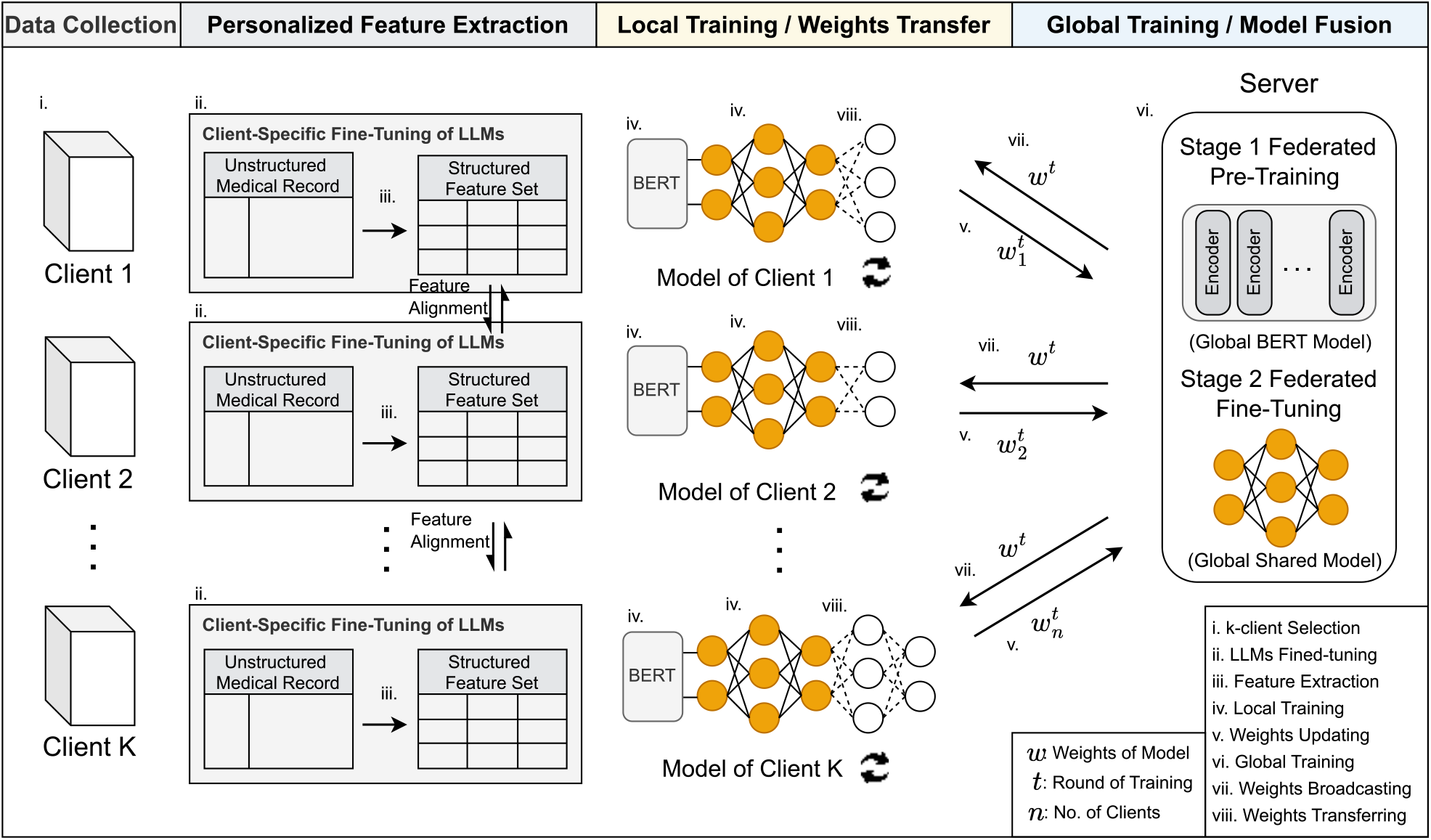
The architecture of Med-SSFWT. The framework operates in several iterative steps: (i) a subset of clients is selected for training in each round; (ii) clients fine-tune pre-trained LLMs on their specific datasets; (iii) structured features are extracted from unstructured medical records; (iv) local models are trained using the extracted features and fine-tuned BERT; (v) clients update their local model weights, which are then; (vi) collected by the server to update the global model; (vii) the updated global model is broadcast back to clients for further local training; and (viii) the process iterates until convergence, achieving a globally optimized model.

As shown in Fig. 3, the federated training of Med-SSFWT operates in two stages: pre-training and fine-tuning. In Stage 1, the BERT model is pre-trained on unstructured text data locally by each client, and the local weights are aggregated by the server to build a global model. In Stage 2, the pre-trained global model is fine-tuned on client-specific tasks, where each client can adopt different task heads, such as MLP, ResNet, or CNNs. Local updates are again aggregated by the server, enabling a globally optimized model while maintaining client personalization. The specific implementation details of Med-SSFWT are presented in Supplementary Alg. 1. The training workflow comprises four key procedures: (1) In Supplementary Proc. 1, each client transforms raw, unstructured medical record data into structured features using fine-tuned large language models, thereby achieving personalized feature extraction. (2) In Supplementary Proc. 2, clients apply information gain-based filtering to retain only those features deemed most informative. (3) In Supplementary Proc. 3, a self-supervised federated pre-training is performed, where the global BERT module is refined by aggregating parameter updates from distributed clients without sharing raw data. (4) Finally, in Supplementary Proc. 4, federated fine-tuning with weight transfer model fusion is conducted, producing in a globally shared model with locally customized client-specific task heads, thereby enabling FTL under strict privacy constraints.

**Figure 3.**
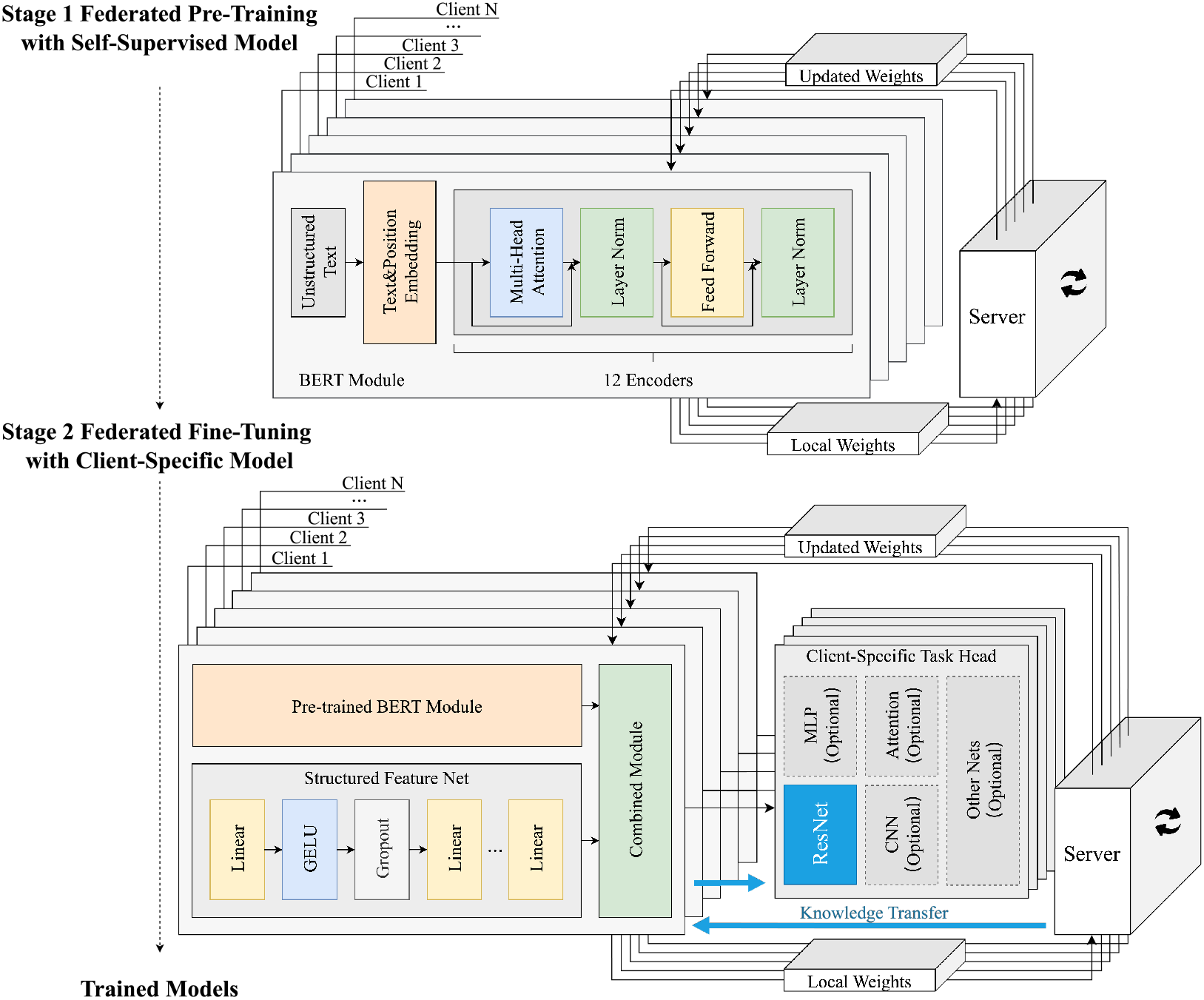
Federated pre-training and fine-tuning stages of Med-SSFWT. In Stage 1, the BERT model is pre-trained on unstructured text data locally by clients using a 12-encoder architecture, including text embedding, multi-head attention, and feed-forward layers. Local weights are aggregated on a central server to update the global model. In Stage 2, the pre-trained model is fine-tuned on client-specific tasks by integrating structured feature networks (e.g., linear layers with GELU) and client-specific task heads, where each client can adopt different network models (e.g., MLP, ResNet, or CNNs) tailored to its specific application. Local updates are aggregated to produce a globally optimized yet personalized model.

### Personalized extraction and alignment of distributed features using LLMs

Each client leverages its local medical dataset and a fine-tuned LLM to transform unstructured medical records into structured feature representations. By maintaining the data and the fine-tuning process entirely on the client side, this step ensures that raw patient data remains strictly localized, thus addressing privacy and regulatory concerns. Specifically, as shown in Supplementary Proc.1, Each client *C*_*i*_ begins by loading its local dataset 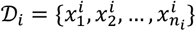, where each 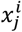 denotes the *j*-th unstructured medical record. A pre-trained language model ℳ_llm_ is fine-tuned on 𝒟_𝒾_ using self-supervised objectives, by minimizing the following loss function, as defined in Eq. (3):

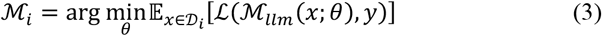

where *θ* denotes the model parameters, and *y* represents auxiliary labels or target outputs derived from self-supervised objectives.

To ensure semantic alignment across clients, a feature-level alignment mechanism is introduced. Each client *C*_*i*_ constructs a local feature schema 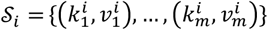, where 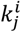 is the feature name and 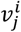 is its associated type or value (e.g., “Smoking History: 40 years, 2 packs/day”, “Body Mass Index: 24.3”, “Kamofsky Performance Status: 80”). Let 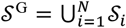 denote the global feature template aggregated across all clients. In the next round, each client performs prompt refinement by aligning its local prompts to match features in 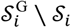, i.e., the set of features discovered from other clients but missing locally. The prompt for record 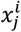 is thus updated as 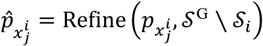. This enables the LLM to extract an augmented feature representation 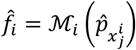 that is both personalized and semantically consistent with peer clients. The final aligned feature set becomes 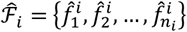. This collaborative feature alignment strategy preserves privacy while improving feature coverage, cross-client consistency, and ultimately, federated model generalizability.

### Information gain-based gradient filtering mechanism

The information gain-based filtering mechanism is designed to selectively integrate client-generated gradient updates into the global model during FL. Instead of indiscriminately aggregating all updates, this mechanism ensures that only those contributing substantive performance improvements are retained by evaluating each gradient update through an adaptive information-theoretic measure. The detailed workflow of IGFM is provided in Supplementary Proc. 2, which describes the gradient evaluation, filtering, and selection procedures integrated into each communication round. Formally, let *θ* be the current set of global model parameters, and consider a client 𝒞_𝒾_ with local dataset 𝒟_𝒾_. Suppose 𝒞_𝒾_ proposes a gradient update ∇*θ*_*i*_, yielding a hypothetical updated parameter set 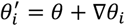. The pre- and post-update expected losses can be defined as 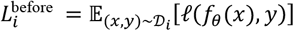 and 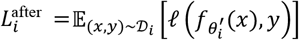, where ℓ(⋅) denotes the loss function and *f*_*θ*_(⋅) represents the multi-task predictive mapping parameterized by *θ*, which jointly predicts the TNM stages from clinical text. The gain in performance contributed by the *i*-th client’s update is quantified as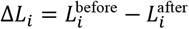.

To ensure stable model training, an information gain criterion is introduced, defined as 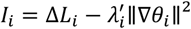, where Δ*L*_*i*_ denotes the loss reduction of client *C*_*i*_ between two consecutive iterations. The adaptive regularization coefficient is expressed as 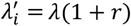, with λ as the base coefficient (set to 0.1 by default) and 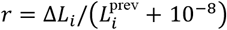 representing the normalized improvement in loss reduction, stabilized by a small constant 10^−8^. A threshold *η* = 0.1 is applied such that only updates satisfying *I*_*i*_ > *η* are accepted, thereby ensuring that the global model evolves based on client contributions that provide substantial and reliable information gain.

### Federated pre-training with self-supervised model

Federated pre-training aims to collaboratively train a global model across multiple distributed clients while preserving data privacy. This process leverages the MLM task to effectively learn semantic representations from unlabeled data within a FL framework. The full training workflow is presented in Supplementary Proc. 3, which outlines the local pre-training steps, update communication, and global aggregation process.

Formally, let ℳ_bert_ represent the global BERT model initialized at the beginning of federated training, and let 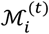 denote the local model for client *i* at training round *t*. During each round, each client optimizes the objective of MLM task on its local dataset 𝒟_*i*_, with the local loss function for client *i* defined in Eq. (4):

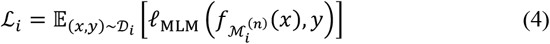

where *ℓ*_MLM_ is the MLM loss, *x* represents the masked input sequence, and *y* represents the target tokens to be predicted.

After completing the local training for a fixed number of epochs (denoted as *E*), the clients send their updated model parameters 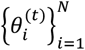 to the central server. The server aggregates these parameters to update the global model 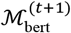 using a weighted averaging strategy, as defined in Eq. (5):

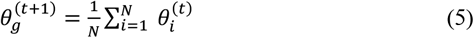

where *N* is the total number of participating clients.

The updated global model 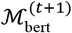 is then distributed back to the clients for the next training round. This iterative process continues until the global model converges or reaches a predefined number of rounds. By leveraging self-supervised learning and federated training principles, this approach enables theglobal model to learn robust representations from distributed medical datasets without compromising data privacy.

### Federated fine-tuning with client-specific model

Med-SSFWT facilitates the application of transfer learning across decentralized clients, each possessing a distinct subset of the data and maintaining personalized client-specific heads tailored to their respective downstream clinical objectives. The shared global model parameters focus on a common feature extraction backbone, while each client keeps its parameters private, avoiding exposing them to the server. Through iterative communication and aggregation, this approach refines the global representation layer, enabling it to better capture commonalities across heterogeneous tasks and data distributions. The complete procedure is detailed in Supplementary Proc. 4, which describes how model fusion is achieved by jointly optimizing the global backbone and local heads.

Consider *N* clients indexed by *i* ∈ {1, …, *N*}. Let *θ*^(H)^ denote the initial global shared parameters and let ϕ_*i*_ represent the local, client-specific parameters associated with unique downstream tasks on client *i*’s side. During each global communication round *t* = {1, …, *T*}, the server broadcasts the current shared parameters *θ*^(*t*−1)^ to all clients. Each client *i* initializes its local model as 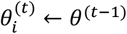 and then locally refines these parameters using 𝒟_*i*_. This local fine-tuning process can be modeled as the minimization of a local objective, as shown in Eq. (6):

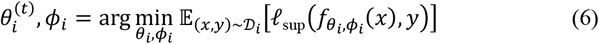

where *ℓ*_sup_(⋅) is a supervised loss function tailored to the downstream task on client *i*, and 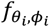 denotes the locally composed model consisting of the shared feature extractor *θ*_*i*_ and the client-specific task head *ϕ*_*i*_.

After local optimization, each client *i* computes the update for the shared parameters, defined as 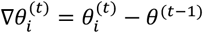, and sends only this gradient update to the server. The private parameters *ϕ*_*i*_ remain local and are not shared or aggregated, ensuring flexibility in downstream tasks and data privacy.

Upon receiving the gradients 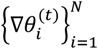, the server applies Supplementary Proc. 2 to determine the subset of clients ℱ^(C)^ ⊆ {1, …, *N*} whose updates are deemed beneficial. This filtering step can be characterized by evaluating an information gain criterion *I*_*i*_ for each client’s gradient and selecting only those with *I*_*i*_ > *η*, where *η* is a predefined threshold.

Let *n*_*i*_ denote the effective number of samples or a weighting factor associated with client *i* ‘s contribution. The server aggregates the filtered gradients as a weighted sum, as shown in Eq. (7):

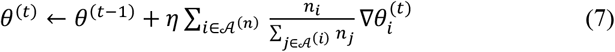

where 𝒜^(*t*)^ represents the set of clients whose updates were accepted after filtering. Over multiple communication rounds, this iterative process leads to a progressively refined global feature extractor *θ*^(*T*)^ that captures robust representations. By separating the global shared parameters from client-specific heads, the paradigm of federated fine-tuning weight transfer model fusion enables crossclient knowledge transfer without forcing identical model architectures. Each client benefits from improved representations learned from diverse data sources, while retaining autonomy and privacy over client-specific capabilities.

### Evaluation metric

To comprehensively evaluate model performance, we employ a range of macro-averaged metrics, including accuracy, precision, recall, and F1-Score. Additionally, convergence-related metrics, such as RoA@σ, AoR@γ, and PCE, are introduced to assess the training dynamics and resource efficiency of models.

The macro-averaged metrics are defined as Eq. (8):

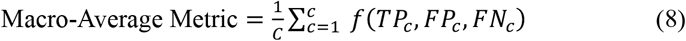

where *C* represents the number of classes, *f*(*TP*_C_, *FP*_C_, *FN*_C_) denotes the specific metric function. Here, *TP*_C_, *FP*_C_, and *FN*_C_ correspond to the True Positives, False Positives, and False Negatives for class *c*, respectively.

RoA@σ measures the number of global rounds required for a FTL model to reach a specified performance threshold σ (e.g., 75% accuracy). It can be defined as Eq. (9):

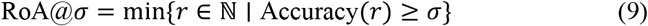

where *r* is the global training round. A smaller RoA indicates faster convergence and higher training efficiency. AoR@ γ quantifies the model’s accuracy at a specified number of global training rounds γ, offering insight into its intermediate performance. It can be defined as Eq. (10):

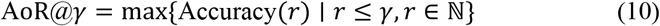

where γ is the specified global round. Higher AoR values indicate better performance within the given rounds, reflecting the model’s effectiveness during the early training. PCE is a comprehensive metric designed to evaluate the efficiency and performance of models under constrained training resources. It combines three key components: AoR@σ, RoA@γ, and the final performance F1-Score. The formulation of PCE is as follows in Eq. (11):

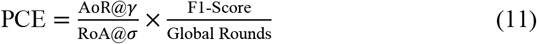

In this experiment, the Dirichlet distribution function is employed to generate datasets with different degrees of non-IID, which is controlled by adjusting the parameter *α* of the distribution^21^. As illustrated in Fig. 4b, different values of *α* yield varying data distributions across clients, where smaller *α* leads to more imbalanced allocations and larger *α* results in more uniform distributions. The Dirichlet distribution is defined as Eq. (12) and Eq. (13):

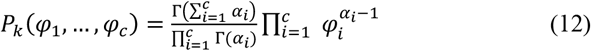

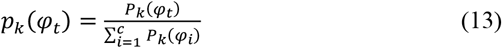

where *φ*_*i*_ represents the proportion of samples in category *i, c* is the total number of categories, and *α* is the parameter of the Dirichlet distribution.

Perplexity is a widely adopted metric for evaluating the quality of language models, particularly in the MLM task^38^. It quantifies the model’s uncertainty in predicting masked tokens and serves as an inverse measure of the model’s confidence in its predictions.

**Figure 4.**
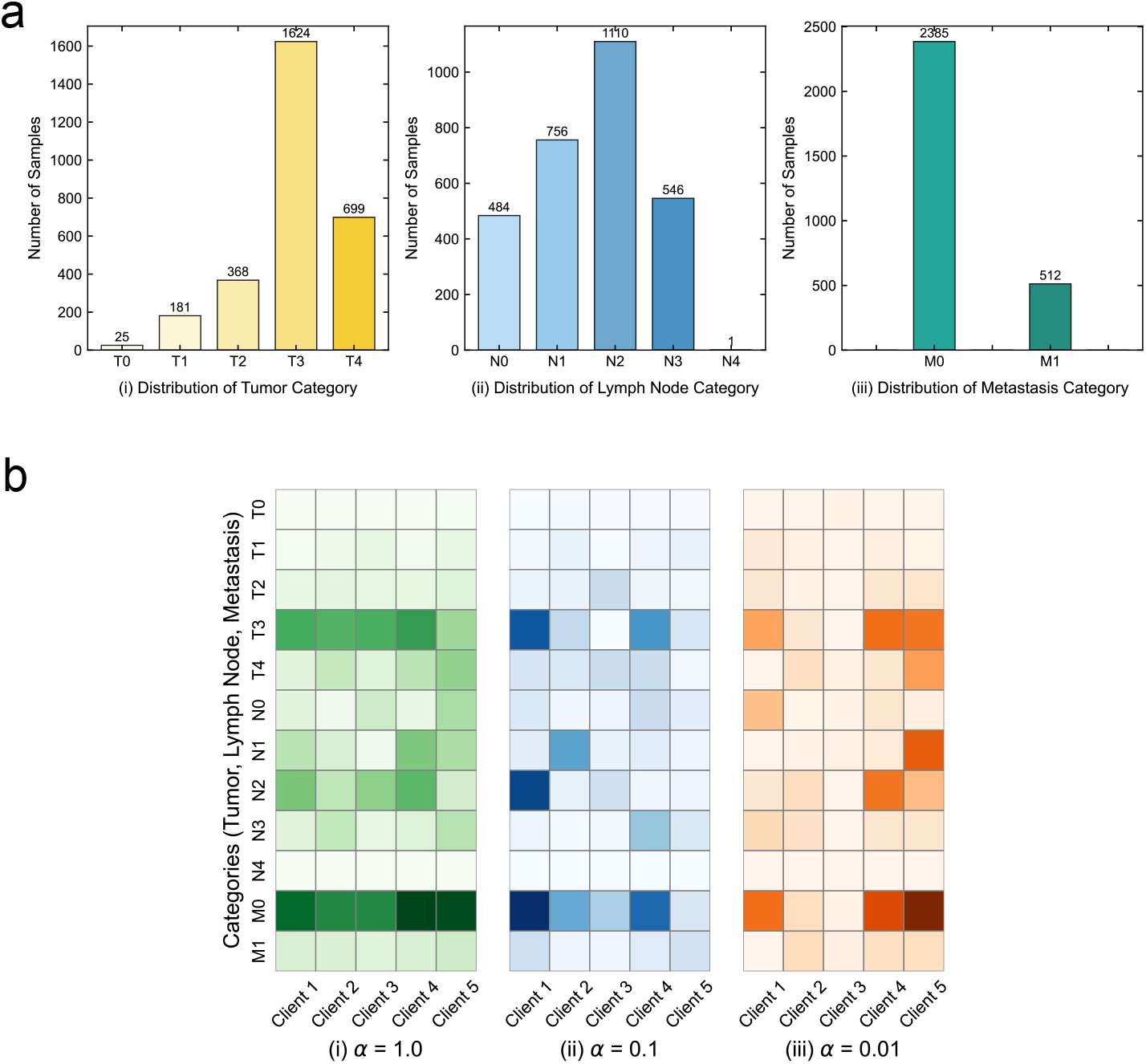
Distribution of TNM stages and data partitioning across clients. (a) Histogram distribution of tumor stages (T0–T4), lymph node involvement (N0– N4), and metastasis status (M0–M1), based on 2,897 labeled samples from the SCCH-ESCA dataset. The data reveal clear imbalances across categories, with tumor stages T3 and T4, lymph node categories N1–N2, and metastasis status M0 dominating the sample distribution. (b) Examples of client-level data allocation under different Dirichlet concentration parameters (*α* = 1.0, 0.1, and 0.01). Higher *α* values correspond to more uniform data distributions across clients, whereas smaller *α* values induce stronger heterogeneity and category imbalance. Darker shades denote larger sample counts for each client-category pair, highlighting the increasing data skew as *α* decreases.

**Figure 5.**
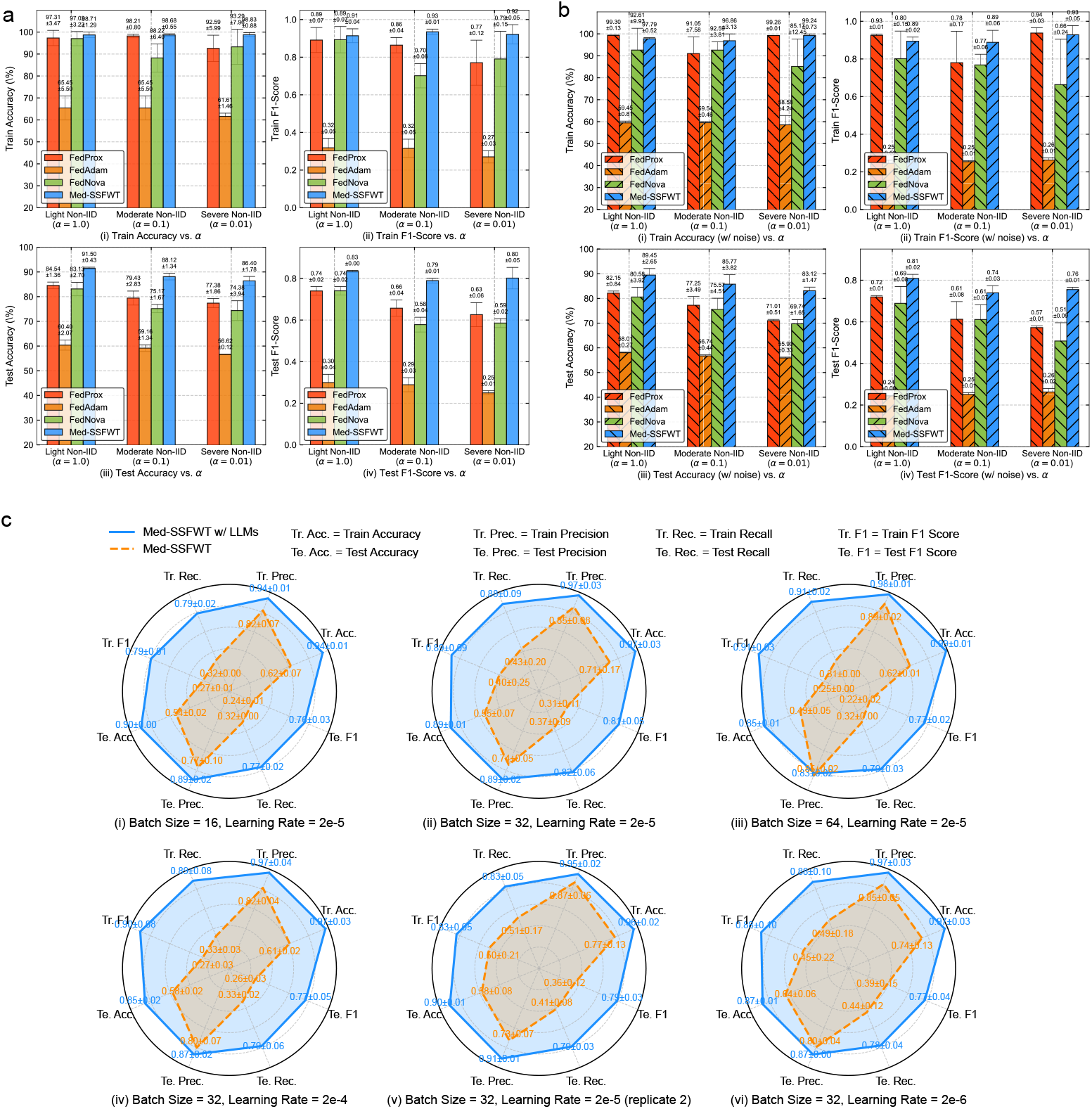
Comparative evaluation under Non-IID conditions and sensitivity of LLM training. (a) Predictive performance of FedProx, FedAdam, FedNova, and Med-SSFWT on TNM staging under Light (*α*=1.0), Moderate (*α*=0.1), and Severe (*α*=0.01) Non-IID settings without additional gradient noise. (b) Predictive performance of the same methods under non-IID settings with additional gradient noise. (c) Evaluation of LLM extraction performance across different batch sizes and learning rates.

**Figure 6.**
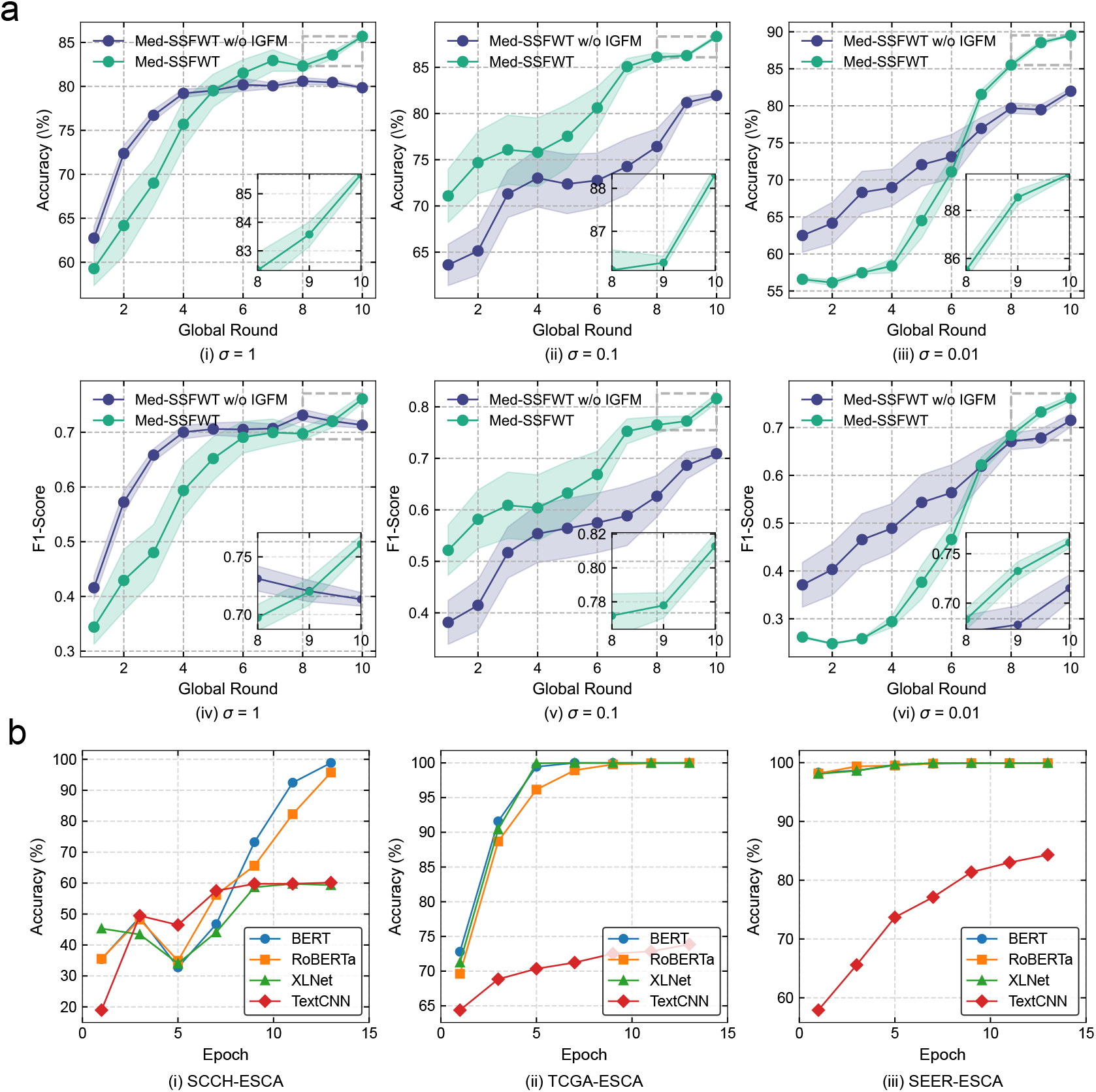
Performance evaluation of gradient filtering in Med-SSFWT and backbone models for the prediction of TNM staging. (a) Effectiveness of the information gain–based gradient filtering mechanism in Med-SSFWT under different gradient noise levels (σ). (b) Evaluation of TextCNN, BERT, RoBERTa, and XLNet backbones for the prediction of TNM staging on the SCCH-ESCA, TCGA-ESCA, and SEER-ESCA datasets.

**Figure 7.**
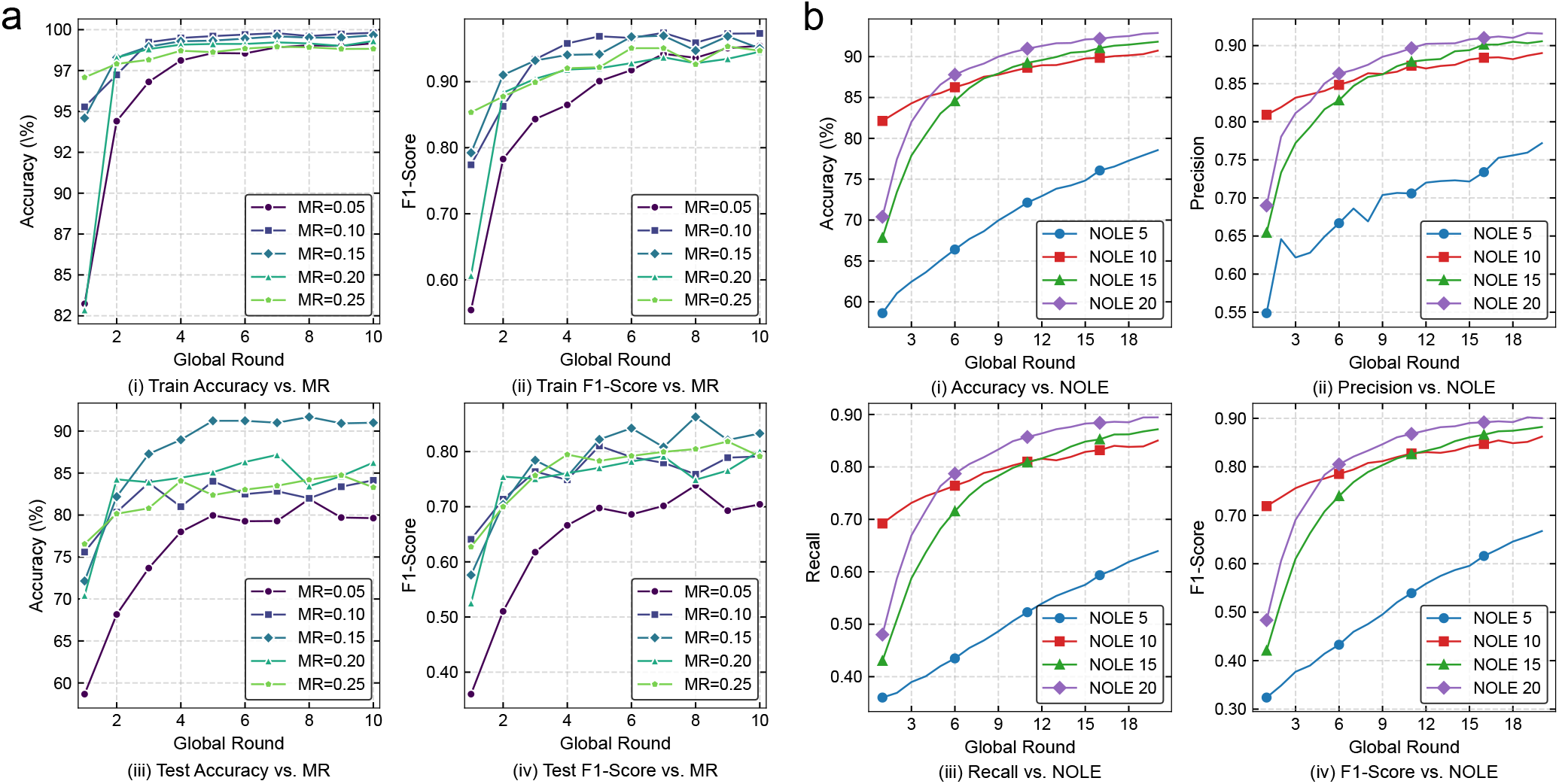
Parameter sensitivity analysis in Med-SSFWT for TNM staging prediction. (a) Comparative training and testing performance under different MR. Impact of the NOLE in Med-SSFWT on the SCCH-ESCA dataset.

Formally, given a sequence of tokens *X* = (*x*_1_, *x*_2_, …, *x*_*n*_), let ℳ_*p*_ denote the masked positions in the sequence, and 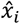 be the model’s predicted probability for the masked token *x*_*i*_ ∈ ℳ_*p*_. The perplexity 𝒫 of the model can be defined as Eq. (14):

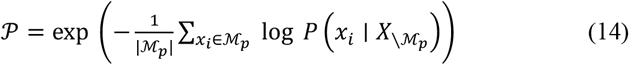

where 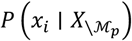 denotes the predicted probability of the masked token *x*_*i*_ conditioned on the unmasked context. A lower value of 𝒫 reflects higher prediction confidence and thus a stronger capacity to model contextual dependencies.

### Ethics Statement

This study was approved by the ethics committee of Sichuan Cancer Hospital (Grant No. KY-2025-337-01) and was conducted in accordance with the Guidelines for Good Clinical Practice and the Declaration of Helsinki^39^. The informed consent requirement was waived by the ethics committee of Sichuan Cancer Hospital due to the retrospective design of the study.

## Supporting information

Supplementary Information for: Med-SSFWT: A Self-supervised Federated Weight Transfer Framework for Medical Model Fusion

## Data Availability

All data produced in the present study are available upon reasonable request to the authors.

## Data availability

The SCCH-ESCA dataset was collected from Sichuan Cancer Hospital under approval of the institutional ethics committee (Grant No. KY-2025-337-01). Due to patient privacy and institutional policies, individual-level clinical records cannot be made publicly available. De-identified data may be accessed for academic research purposes upon reasonable request to Sichuan Cancer Hospital and subject to a data use agreement.

The TCGA-ESCA dataset is publicly available from The Cancer Genome Atlas (https://portal.gdc.cancer.gov). The SEER-ESCA dataset is publicly available through the Surveillance, Epidemiology, and End Results program of the U.S. National Cancer Institute (https://seer.cancer.gov).

All other relevant data supporting the findings of this study are available within the paper and its Supplementary Information files. Additional data are available from the corresponding author upon reasonable request.

## Code availability

The source code implementing Med-SSFWT, including preprocessing scripts, model training and evaluation routines, and parameter configurations, is available at https://github.com/MaoLab-CD/Med-SSFWT.

## Acknowledgments

This work was supported by the National Natural Science Foundation of China (No. T2322002), and the Sichuan Science and Technology Program (No. 2024NSFTD0032, 2024ZYD0006, 2025ZNSFSC0003) to X.W.M.; and by the Sichuan Science and Technology Program (No. 2025ZNSFSC1094), the Noncommunicable Chronic Diseases-National Science and Technology Major Project (No. 2023ZD0502300), the Postdoctoral Fellowship Program (Grade C) of China Postdoctoral Science Foundation (No. GZC20251741) to Y.J.H. The authors acknowledge the support of the Center for HPC at the University of Electronic Science and Technology of China.

## Author Contributions

Q.H., Y.H., and K.Z. conceived this idea. Q.H. and X.M. designed and supervised the project. Q.H., Y.H., K.Z., Q.T., and X.M. drafted the manuscript. R.Y., Z.Z., Y.X., and Z.W. performed the experiments and data analysis. Q.H. prepared the figures. K.Z., S.Z.M.I., I.M.M., Y.Z., Q.Y., H.Z., and D.W. contributed to data collection, clinical interpretation, and critical revision of the manuscript. All authors read and approved the final version of the manuscript.

