## Supplementary Information for: Med-SSFWT: A Self-supervised Federated Weight Transfer Framework for Medical Model Fusion for "Med-SSFWT: A Self-supervised Federated Weight Transfer Framework for Medical Model Fusion"

### Supplementary Materials

This Supplementary Information provides additional technical details, experimental visualizations, and algorithmic procedures supporting the main text. Supplementary Figures illustrate the empirical behaviors and performance trends of the proposed Med-SSFWT framework, providing complementary evidence to the quantitative results reported in the main text. Supplementary Algorithm 1 then presents the complete implementation of the framework, including four key procedures: (1) personalized extraction of distributed features using fine-tuned LLMs; (2) information gain-based adaptive gradient filtering; (3) federated pre-training with self-supervised BERT; and (4) federated fine-tuning with weight transfer model fusion.

#### Supplementary Figures

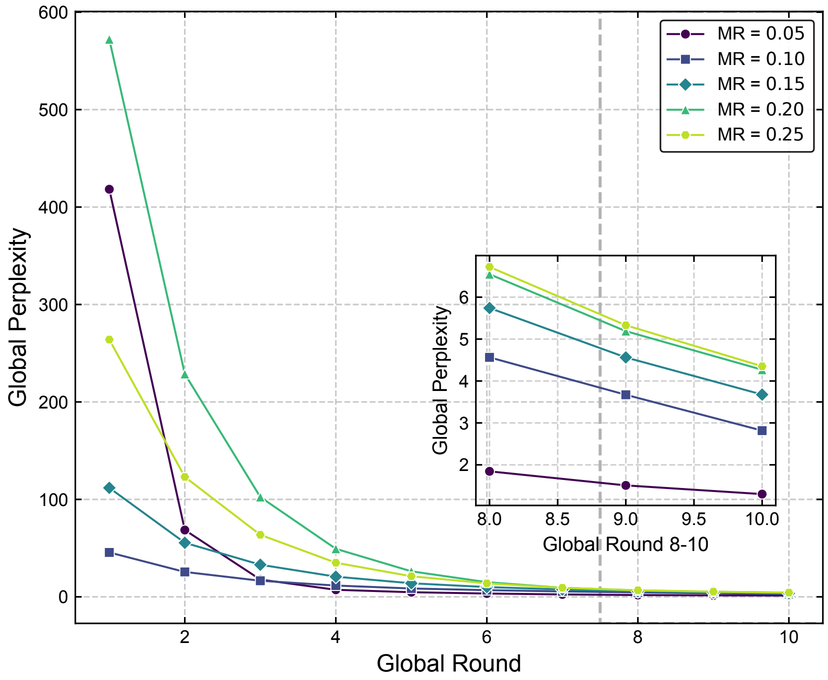


**Figure 1:** Comparative analysis of global perplexity reduction during the self-supervised pre-training stage of Med-SSFWT under different masking ratios (MR = 0.05, 0.10, 0.15, 0.20, and 0.25). The y-axis denotes the global perplexity values aggregated across federated clients, while the x-axis shows the number of global communication rounds (from 1 to 10). Across all configurations, global perplexity decreases rapidly in the early rounds and stabilizes at low values by the 6th-8th round, indicating effective convergence. Notably, lower masking ratios (MR = 0.05 and 0.10) achieve faster and lower perplexity convergence, whereas higher ratios (MR = 0.20 and 0.25) converge more slowly and remain at higher perplexity values. The inset highlights the differences among masking ratios in the later stages (rounds 810), showing that MR = 0.15 achieves a balanced trade-off between convergence speed and final perplexity, consistent with its superior downstream performance.

### Supplementary Algorithm

**Algorithm 1:** The implementation of Med-SSFWT

**Input:** datasets of medical data $\{\mathcal{D}_1, \mathcal{D}_2, \ldots, \mathcal{D}_n\}$

**Output:** shared global model $\mathcal{M}^*$, personalized local model $\{\mathcal{M}_1^*, \mathcal{M}_2^*, \ldots, \mathcal{M}_n^*\}$

Each client $\mathcal{C}_i$ load $\mathcal{D}_i$;

Execute **Proc. 1** *Personalized Extraction and Alignment of Distributed Features*:

Fine-tune $\mathcal{M}_{llm}$ on $\mathcal{D}_i$ to obtain $\mathcal{M}_i$, extract structured features $\mathcal{F}_i$ from prompts, derive local schema $\mathcal{S}_i$ and exchange with peers to construct global schema $\mathcal{S}_G$, then identify missing fields $\mathcal{S}_i^{\text{missing}}$ and regenerate aligned feature set $\mathcal{F}_i'$;

Execute **Proc. 2** *Information Gain-based Gradient Filtering*:

Each model $\mathcal{M}_i$ calculates information gain for each feature in its local dataset and filters out low-relevance features based on a predefined threshold $\tau$;

Execute **Proc. 3** *Federated Pre-training with Self-supervised BERT*:

$\mathcal{M}_{bert}^* \leftarrow$ Federated pre-taining the global BERT module $\mathcal{M}_{bert}$ via parameter averaging under a self-supervised objective across clients;

Execute **Proc. 4** *Federated Fine-tuning with Weight Transfer Model Fusion*:

$\mathcal{M}^*, \{\mathcal{M}_1^*, \mathcal{M}_2^*, \ldots, \mathcal{M}_n^*\} \leftarrow$ Federated fine-tuning the shared global model $\mathcal{M}_{shared}$ and locally fine-tune $\mathcal{M}_i$'s task head with its private dataset $\mathcal{D}_i$. ;

**return** $\mathcal{M}^*, \{\mathcal{M}_1^*, \mathcal{M}_2^*, \ldots, \mathcal{M}_N^*\}$;

**Procedure 1:** *Personalized Extraction and Alignment of Distributed Features*

**Input:** Distributed medical datasets $\{\mathcal{D}_1, \ldots, \mathcal{D}_N\}$, a pre-trained LLM $\mathcal{M}_{llm}$.

**Output:** Aligned feature sets $\{\hat{\mathcal{F}}_1, \ldots, \hat{\mathcal{F}}_N\}$ for all clients.

```
foreach client i = 1, ..., N do
    D_i ← Load Local Dataset;
    M_i ← FineTune(M_llm, D_i);
    S_i ← ∅
    foreach record x ∈ D_i do
        p_x ← ConstructPrompt(x);
        f_x ← M_i(p_x);
        F_i ← F_i ∪ {f_x};
        S_i ← S_i ∪ ExtractFeatureSchema(f_x);
    end
end
S^G ← ⋃_{i=1}^{N} S_i ;
foreach client i = 1, ..., N do
    S_i^missing ← S^G \ S_i;
    Update prompt strategy to include S_i^missing;
    foreach record x ∈ D_i do
        p̂_x ← Refine(x, S_i^missing);
        f̂_x ← M_i(p̂_x);
        F̂_i ← F_i ∪ {f̂_x};
    end
end
return {F̂_1, ..., F̂_N}
```

**Procedure 2:** *Information Gain-based Gradient Filtering*

**Input:** Client gradients $\{\nabla\theta_i\}_{i=1}^{N}$, Global model parameters $\theta$, Client data distributions $\{\mathcal{D}_i\}_{i=1}^{N}$

**Output:** Filtered gradients $\{\nabla\theta_i\}_{i\in\mathcal{A}}$

Initialize threshold $\eta$;

Initialize regularization parameter $\lambda$;

Initialize accepted set $\mathcal{A} \leftarrow \emptyset$;

**for** $i \leftarrow 1$ **to** $N$ **do**

  $\theta_i' \leftarrow \theta + \nabla\theta_i$;

  $L_i^{\text{before}} = \mathbb{E}_{(x,y)\sim\mathcal{D}_i}\left[\ell\left(f_\theta(x), y\right)\right]$;

  $L_i^{\text{after}} = \mathbb{E}_{(x,y)\sim\mathcal{D}_i}\left[\ell\left(f_{\theta_i'}(x), y\right)\right]$;

  $\Delta L_i = L_i^{\text{before}} - L_i^{\text{after}}$;

  $r_i = \frac{\Delta L_i}{L_i^{\text{before}}+\epsilon}$;

  $\lambda_i' = \lambda(1 + r_i)$;

  $\|\nabla\theta_i\| = \sqrt{\sum_{j=1}^{d}\left(\nabla\theta_i^{(j)}\right)^2}$;

  $\mathcal{I}_i = \Delta L_i - \lambda_i'\|\nabla\theta_i\|^2$;

  **if** $\mathcal{I}_i > \eta$ **then**

    $\mathcal{A} \leftarrow \mathcal{A} \cup \{i\}$;

  **end**

**end**

**return** $\{\nabla\theta_i\}_{i\in\mathcal{A}}$

**Procedure 3:** *Federated Pre-training with Self-supervised BERT*

**Input:** Global BERT model $\mathcal{M}_{bert}$, number of global rounds $T$, local epochs $E$, learning rate $\eta$, client datasets $\{\mathcal{D}_i\}_{i=1}^N$

**Output:** Pretrained global BERT model $\mathcal{M}_{bert}^*$

```
Initialize global model parameters θ_g;
for each global round t = 1, ..., T do
    Broadcast global model M_bert with parameters θ_g to all clients;
    Initialize client models {M_i}_{i=1}^N with θ_g;
    Client updates:
    for each client i = 1, ..., N in parallel do
        Initialize local model M_i with θ_g;
        for each local epoch e = 1, ..., E do
            for each batch (x, y) ∈ D_i do
                Mask input x to generate x_masked;
                Compute MLM loss ℓ_MLM(f_{M_i}(x_masked), y);
                Update local parameters θ_i ← θ_i − η∇_{θ_i} ℓ_MLM;
            end
        end
        Send updated local parameters θ_i to the server;
    end
    Server updates:
    Aggregate local parameters, θ_g ← (1/N) Σ_{i=1}^N θ_i,
    Update global model M_bert with θ_g;
end
return M*_bert with parameters θ_g
```

**Procedure 4:** *Federated Fine-tuning with Weight Transfer Model Fusion*

**Input:** Initial global shared parameters $\theta^{(0)}$, Number of communication rounds $T$, Set of clients $\mathcal{C} = \{1, 2, \ldots, N\}$, Learning rate $\eta$, Labeled datasets $\{\mathcal{D}_i\}$ for local fine-tuning

**Output:** Trained global shared parameters $\theta^{(T)}$

Initialize global shared parameters $\theta^{(0)}$;

**for** *each global round* $t = 1, \ldots, T$ **do**

  Broadcast global model $\mathcal{M}_{share}$ with parameters $\theta^{(t-1)}$ to all clients;

  **Client Updates:**

  **for** *each client* $i \in \mathcal{C}$ ***in parallel*** **do**

    Initialize local copy of shared parameters: $\theta_i^{(t)} \leftarrow \theta^{(t-1)}$;

    Let $\phi_i$ represent client $i$'s specific local model parameters (not shared);

    $(\theta_i^{(t)}, \phi_i) \leftarrow \text{LocalFineTune}(\theta_i^{(t)}, \phi_i; \mathcal{D}_i, \eta)$;

    $\nabla\theta_i^{(t)} \leftarrow \theta_i^{(t)} - \theta^{(t-1)}$;

    Send $\nabla\theta_i^{(t)}$ to server;

  **end**

  **Server Updates:**

  Apply Proc. 2 *Information Gain-based Gradient Filtering* to $\{\nabla\theta_i^{(t)}\}_{i=1}^{N}$ to obtain accepted set $\mathcal{A}^{(t)}$;

  $\theta^{(t)} \leftarrow \theta^{(t-1)} + \eta \sum_{i \in \mathcal{A}^{(t)}} \frac{n_i}{n_{\mathcal{A}}} \nabla\theta_i^{(t)}$, where $n_{\mathcal{A}} = \sum_{i \in \mathcal{A}^{(t)}} n_i$;

**end**

**return** $\theta^{(T)}$
